# Small-form-factor LED-based light application surface for indocyanine green-mediated antibacterial photothermal therapy

**DOI:** 10.1101/2020.09.08.20187161

**Authors:** Sakari Nikinmaa, Niina Moilanen, Timo Sorsa, Juha Rantala, Heikki Alapulli, Anja Kotiranta, Petri Auvinen, Esko Kankuri, Jukka Meurman, Tommi Pätilä

## Abstract

We present here a small-form-factor LED-based antimicrobial photothermal therapy (aPTT) applicator surface, and its clinical feasibility as well as efficacy for administration of indocyanine green (ICG)-assisted aPTT to the dental plaque.

Fifteen healthy adults were assigned to this four-day randomized study. After rinsing with ICG, 100J/cm^2^ of 810 nm light was applied to the aPTT-treatment area. Plaque area and gingival crevicular fluid (GCF) matrix metalloproteinase-8 (MMP-8) were measured, and plaque bacteriomes before and after the study were analyzed using 16S rRNA sequencing.

aPTT administration was preformed successfully and plaque-specifically with the applicator surface. Total plaque area and endpoint MMP-8 levels were reduced on the aPTT-treatment side. aPTT reduced *Streptococcus, Acinetobacteria, Capnocytophaga*, and *Rothia* bacteria species in plaques. The applicator surface is feasible for aPTT self-administration, targets therapy to dental plaque, reduces plaque forming bacteria and exerts anti-inflammatory and –proteolytic effects.

## INTRODUCTION

Dental diseases affect a staggering 3.5 billion people globally.^[1,2]^ Dental caries or tooth decay is the most common worldwide health condition, while gingivitis, gum inflammation, affects 50–90% of adults.^[1,2]^If untreated, gingivitis can lead to periodontitis, a more severe inflammation causing irreversible oral tissue destruction and bone resorption around the teeth. Severe periodontitis, the major cause for tooth loss,^[3-5]^ has a prevalence of 5–15% in most populations.^[1,2]^ Importantly, poor oral hygiene and oral microbial dysbiosis are associated with the development or worsening of the course of several systemic diseases, thus adversely affecting the entire body.^[5]^

Antimicrobial photothermal therapy (aPTT) uses light emission to excite photosensitizer molecules. The resulting release of heat energy, and to a lesser degree reactive oxygen species (ROS), destroy bacteria, fungi and viruses.^[6,7]^ Phototherapy, in general, is used in medicine to treat a wide range of medical conditions, including acne, psoriasis, and other skin conditions as well as different types of cancer.^[8]^

Indocyanine green (ICG) is a US Food and Drug Administration-approved dye for medical purposes. ICG is primarily employed in diagnostic approaches, such as assessment of hepatic function and perfusion-related analysis of tissues, due to its non-toxic nature and fast metabolism.^[9]^ Recently, owing to its absorption peak close to the 810nm wavelength of dental caries removal lasers, ICG has arisen as a promising tool in dentistry. Upon excitation, ICG transforms approximately 88–85% of the absorbed energy to heat while less than 15% is released as singlet oxygen.^[10]^ ICG-mediated aPTT has demonstrated therapeutic potential on dental pathogens and periodontitis.^[10-16]^

The administration of laser-assisted aPTT is limited to in-office use by dental professionals. In order to obtain most benefit, aPTT would require routine daily administrations, preferably as self-care.^[17]^ Moreover, a small self-care instrument would facilitate oral health not only in high-income countries but also in low- and middle-income countries in which access to preventive oral healthcare and treatments can be limited.^[18]^

We present here an LED-based light application surface designed for self-managed aPTT administration (**Figure 1A**), and report its feasibility and efficacy in a randomized, split-mouth treatment-control clinical study on 15 healthy volunteers. We demonstrate the applicator surface’s clinical feasibility as well as efficacy to decrease plaque formation, reduce the levels of gingivitis- and periodontitis-associated matrix metalloproteinase-8 (MMP-8) and reduce the number of cariogenic streptococci in dental plaque. Our results demonstrate the feasibility of the small-scale material design for self- or home-application and provide the first evidence to support its efficacy to reduce the amount of dental plaque.

**Figure 1.**
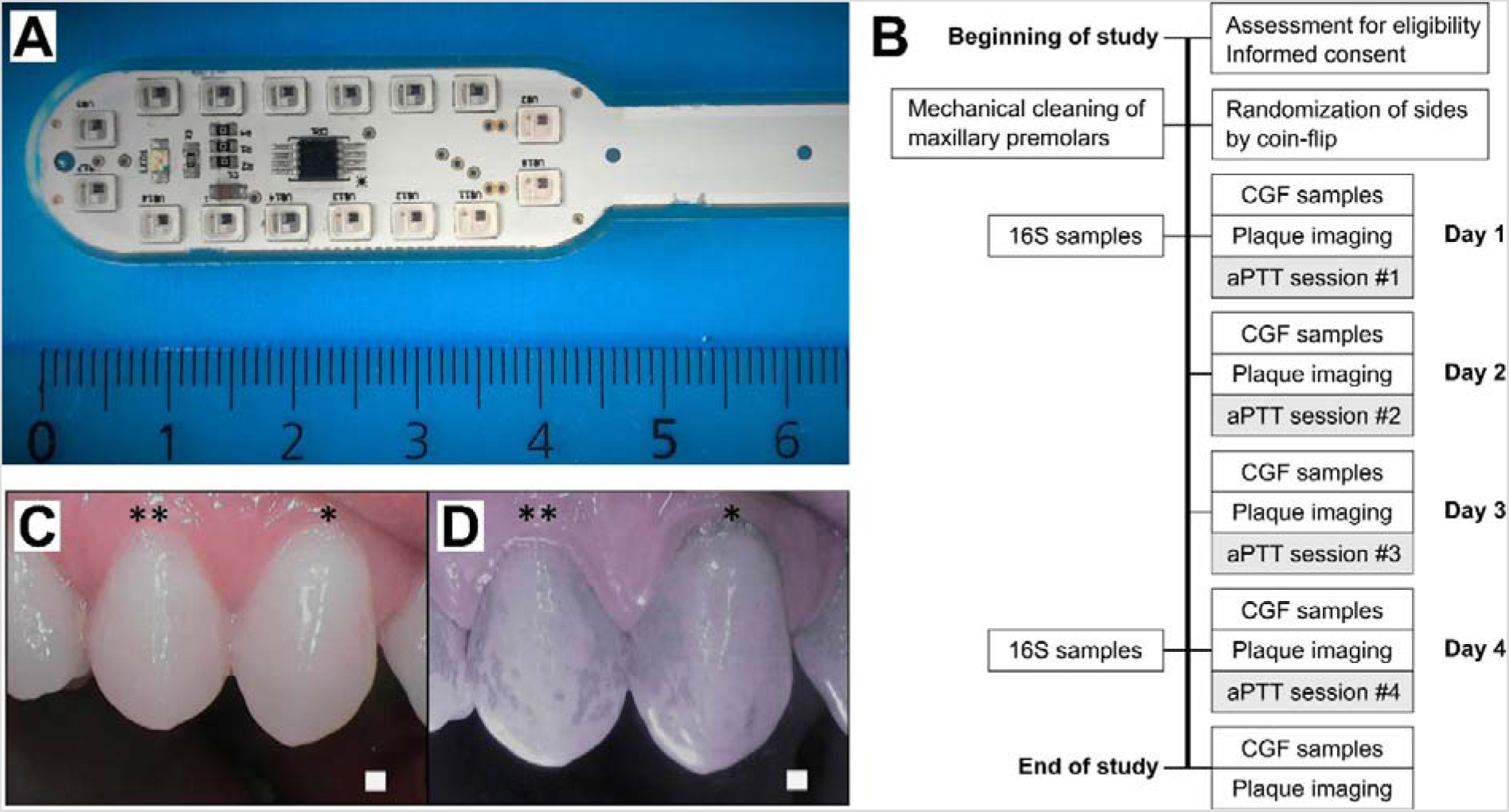
**A.** The small-form-factor LED-based light applicator surface in lollipop form. The linear scale is in centimeters **B**. Study workflow. 16S, bacteriome 16S rRNA sequencing; CGF cervical gingival fluid; aPTT, antimicrobial photothermal therapy. **C, D**. Selective indocyanine green (ICG) localization to the dental plaque. Daylight (**C**) and (**D**) near-infrared light images after ICG mouth rinse. Representative images at day 2, when the teeth had been without cleaning for a single day. * premolar one and ** premolar two.

## MATERIALS AND METHODS

The study was approved by the ethics committee of the Hospital District of Helsinki and Uusimaa (HUS/827/2018) and was conducted in accordance with the ethical principles of the Declaration of Helsinki. All participants provided written informed consent before enrolment. The study is registered in the ISRCTN registry (http://www.isrctn.com under the trial number ISRCTN36318197. All variables were analyzed blinded to the treatment allocation.

### Study design

This randomized, split-mouth study included 15 healthy volunteers. **Figure 1B** presents the study design. aPTT was administered for a total of four times on four consecutive days using the small-form-factor LED-based light applicator surface (**Figure 1A**) after spurting the ICG photoactive substance. Inclusion criteria included good general health, age 18 to 65, and ability to refrain from brushing one’s teeth throughout the study period. Exclusion criteria were diabetes, medications that may affect the immune response or saliva secretion, malignancy, pregnancy, dental implants or prosthesis, fixed orthodontic appliances, or active or chronic oral infection. Medications that could affect the oral microbiome, such as antibiotics, chlorhexidine or other antimicrobial mouthwashes, were not permitted during the study.

Two days before the study, a meticulous professional cleaning was performed on maxillary first premolars on both sides. The treatment side was randomized by coin flip, and the contralateral side served as the control. Study subjects were not allowed to brush or otherwise clean their teeth during the study period.

### aPTT application

ICG powder (Verdye, Diagnostic Green GmBH, Aschheim, Germany) was dissolved in water at a weight/volume ratio of 7 mg/25 ml, and each subject spurted the solution for 60 s in their mouth prior to the light application. Near-infrared imaging was performed to ensure the localization of the ICG in the dental plaque (**Figure 1C,D**).

The small-form-factor LED-based light applicator surface included 16 0.5 W LEDs in a lollipop form, optimally located for producing an even light distribution (**Figure 1A**) to the teeth’s surface.

Light application time was determined by the target light dose of 100 J/cm^2^, and the treatment time was set accordingly for 8 minutes per session. Light intensity was decreased if the person under examination felt that the heat of the light applicator was too intense. In such case, the reduction in light intensity was adjusted by an increase in treatment time to regulate the target dose. This adjustment was performed with a light power meter (Thorlabs PM 100D with S121C sensor head, Thorlabs Inc, Newton, NJ, USA).

### Plaque analysis

The premolar teeth were photographed daily on both sides of the maxilla using a ProDENT PD740 dental camera (Venoka USA Inc, Windermere, FL, USA) under near infrared and white light lighting conditions to assess ICG adherence to the dental plaque. Daily imaging using a SoproCare camera (Acteon Group Ltd, Norwich, England) was carried out to assess the development of plaque formation. The final plaque imaging was performed with the ProDENT PD740 camera after plaque staining with an erythrosine tablet according to the manufacturer’s instructions. The amount of dental plaque surface area was measured from the images of both adjacent upper premolars using Photoshop CC software (Adobe Inc, San Jose, CA, USA). On each side, the amount of plaque was determined as the plaque area pixels divided by the total maxillary premolar teeth area pixels.

### GCF collection and MMP-8 analysis

GCF samples were collected from the first premolar teeth on each side of the maxilla. Collection of GCF and measurement of clinical parameters were performed prior to any treatment measures, daily before the treatment, and after the last treatment. A total of 72×2 GCF samples were collected from the treatment and control sites. GCF sampling was performed by inserting a PerioPaper strip (Oraflow Inc, NY US) into the orifice of the gingival sulcus. Samples were collected at the buccal surface, with the insertion point only minorly changing with every sample. GCF sampling was done carefully and it did not remove any observable amount of supra gingival plaque Strips contaminated by blood were discarded. The samples were stored in small aliquot containers and kept at –20° C until analysis. MMP-8 levels were determined by a time-resolved immunofluorescence assay (IFMA) as described previously.^[19-22]^.

### 16S rRNA bacterial samples

Plaque samples were obtained with Iso Taper Paper Points, size-20 (VDW GmbH, Munich, Germany), by scrubbing the plaque on the tooth enamel. In all the plaque samples, the plaque was intact at the gingival boundary. Each sample was taken from same site by horizontally placing the paper point above gumline of premolar teeth. The paper points were placed into sterile, small-aliquot containers, and were immediately stored at –20°C until analysis.

16S rRNA sequencing was performed as described earlier.^[23]^ The V3-V4 regions of the 16S rRNA genes were amplified using universal bacterial primers and sequenced with an Illumina MiSeq sequencer (Illumina Inc., San Diego, CA, USA). Bioinformatic analysis, including operational taxonomic unit (OTU) clustering and taxonomy assignment were done using Mothur software.^[24]^ The 16S rRNA sequences have been deposited to the Sequence Read Archive (SRA) of the National Center for Biotechnology Information (NCBI) under BioProject ID PRJNA661546.

### Statistical analysis

Statistical analyses were performed using GraphPad Prism 6.0 (GraphPad Software Inc., San Diego, CA, USA). Unpaired comparisons between the groups were performed with the Mann-Whitney test. The paired rank-sum Wilcoxon test was used for the paired samples. Two-way Anova was used to compare the daily values of MMP-8 between groups. Comparisons of categorical variables were performed using Fisher’s exact test. Data are presented as mean ±SEM unless otherwise specified.

## RESULTS

Thirteen subjects completed the entire study protocol. One dropout occurred two days into the study due to a non-related acute infection, and another four days into the study, just after the 16S sampling and just before the plaque imaging, due to a work-related emergency.

### Dental plaque formation

All subjects were plaque-free at the beginning of the study period. ICG was demonstrated to attach selectively to the dental plaque (**Figure 1C**). Sopro chromatic mapping did not show any difference in the development speed of the plaque between the aPTT-treatment and control sides. At the end of the study, erythrosine-based plaque-enclosing imaging showed significantly less plaque on the aPTT-treated premolars than controls(35.1±4.8% vs 42.5± 4.0% of the surface area; p = 0.016; **Figure 2**).

**Figure 2.**
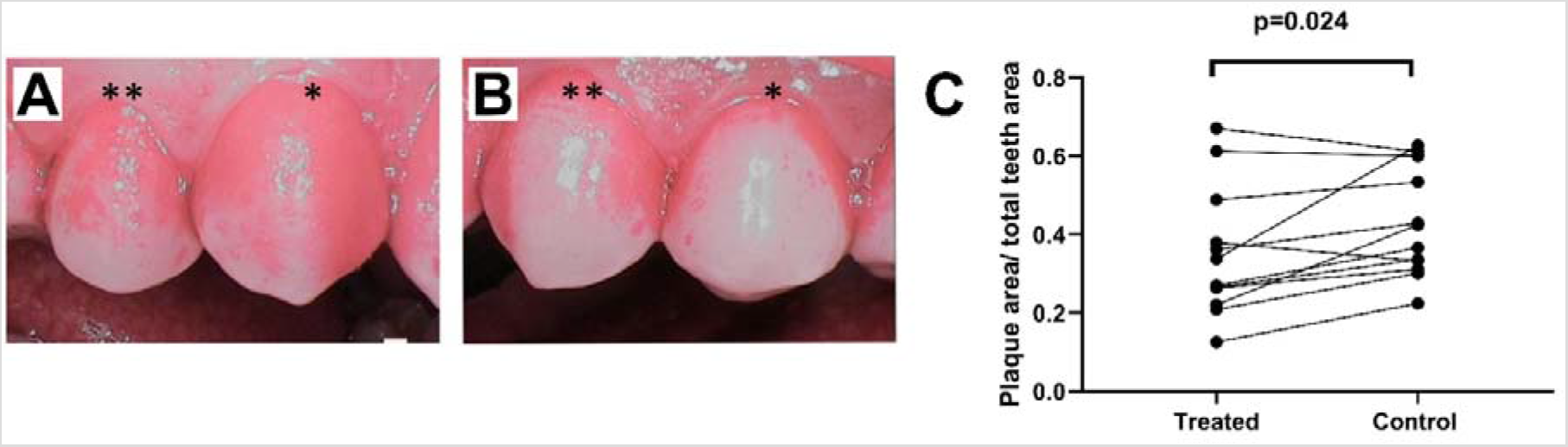
**A,B.**Comparison of dental plaque formation. Panel **A** shows plaque (area 62.7% of total premolar dental area) on the control side in the last imaging session. Premolar one (*) and premolar two (*). Panel **B** shows plaque (area 33.8% of total premolar dental area) on the treatment side at the same time point. **C**. Plaque areas at the end of study after four days of aPTT application. Paired measurements demonstrated significantly less plaque formation on the treated side compared to control side.

### Gingival crevicular fluid MMP-8 levels

At the beginning of the study, two days after meticulous professional cleaning GCF MMP-8 concentrations on both sides were similar. Evaluation of daily pre-treatment MMP-8 concentrations throughout the study using an area under curve analysis demonstrated a trend for lower overall MMP-8 concentrations in the treated group (AUC 256.7+99.7, 95%CI 61.2–452.1) as compared to the control side (AUC 344.9+113.2, 95%CI 123.0–566.8). However, the GCF MMP-8 levels at the end of the study after aPTT administration demonstrated a significant reduction on the aPTT-treated side (**Figure 3A**).

**Figure 3.**
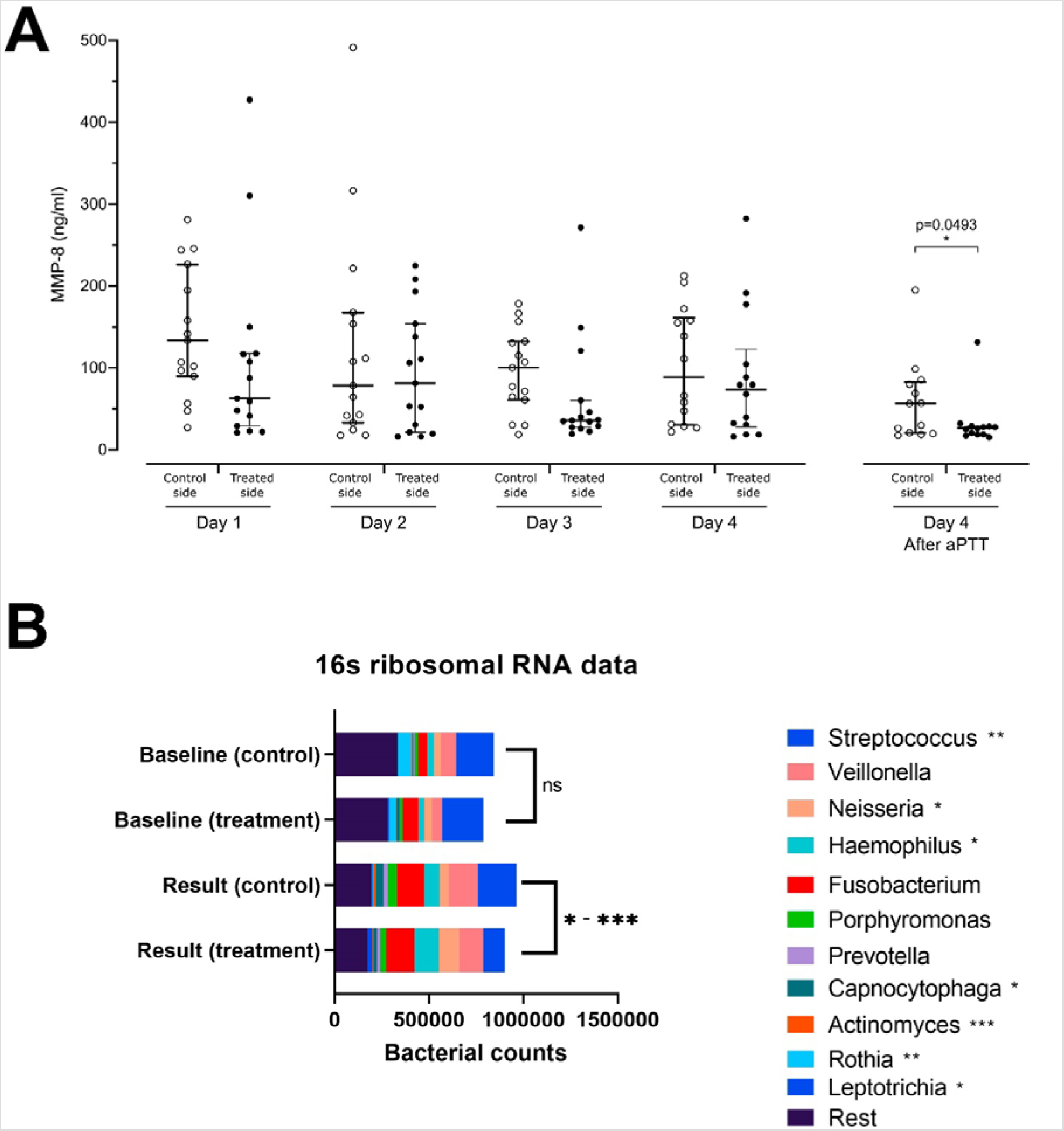
A.Matrix metalloproteinase-8 (MMP-8) concentrations at indicated days and sides in samples from gingival pockets. aPTT effect on MMP-8 secretion significantly lower on the aPTT-treated side than on the control side after the last application of aPTT at end-of-study. **B**. Analysis of bacteriome using 16S rRNA sequencing. This figure shows the diversity of bacteria within the plaque samples from the treatment and control premolars at the start of the treatment at baseline and at the end of the study period. At the end of treatment, a significant reduction in the relative proportion of *Streptococcus, Actinomyces*, and *Rothia* bacteria species was identified, and a relative increase in the *Neisseria, Haemophilus*, and *Leptotrichia* bacteria species was seen between treated and control side. * p< 0.05, **p< 0.01, ***p< 0.001

### Microbiome analysis and alpha diversity

Fourteen paired pre-treatment and posttreatment 16S samples were collected. One sample was disqualified due to low sample quality, because it showed significantly lower bacterial counts together with a significantly reduced amount of OTUs detected. The number of streptococcal species on the aPTT-treated side (median 7460, range 6734–23987) was significantly reduced as compared to the control side (median 12776, range 4959–38669, p = 0.0024). Similarly, reduced numbers of Rothia species (median 13, range 3–908 vs. median 82, range 3–2883, p = 0.0032), Capnocytophaga species (median 1050, range 11–4382 vs. median 1220, range 15–8599; p = 0.0402) and Actinomyces species (median 141, range 16–1833 vs. median 933, range 12–3608, p = 0.0005) were detected on the aPTT-treated vs control sides. Increases in the OTU-amounts of Neisseria [median 7211, range 201–22892 vs. median 2169, range 122–11792, p = 0.0327), Haemophilus species [median 7599, range 713–28727 vs. median 2935, range 524–27988, p = 0.0479), and Leptotrichia (median 307, range 1–5524 vs. median 105, range 4–3431, p = 0.0105) were found on the aPTT-treated vs. the control side(**Figure 3B**).

## DISCUSSION

We present here a surface assembly for self-administration of aPTT and demonstrate its feasibility and effectivity in a four-day clinical study. In addition to self-administration feasibility, our results demonstrated reduced dental plaque, reduced the levels of the gingivitis and periodontitis biomarker GCF MMP-8 as well as reduced amount of cariogenic and periodontitis-associated bacteria on the aPTT-treated side. Moreover, alpha diversity of the plaque microbiome was retained throughout the study period.

Plaque-selective binding of ICG photosensitizer occurred effectively after solution spurting suggesting a targeted effect on the plaque bacteria sensitive to aPTT.^[25]^ Further targeting is achieved through localized ICG excitation by the LED-based light applicator surface. In contrast to the coherent and intense laser beam used in laser-based devices, the LED light sources produce scattered, wide-spreading light without hotspots. This reduces risks associated with laser light, such as eye damage and thermal injury to tissues.^[26]^ An additional benefit of the LED-based surface is the improved total amount of light when compared to laser devices. Finally, the most enticing element of LED light is the ease of use and general safety, which are important for daily use at home.

In samples collected daily before aPTT administration, no significant overall differences in GCF MMP-8 were observed during the study period. However, when collected after the administration of aPTT at the end of the study, the levels of GCF MMP-8 were significantly lower on the treated side as compared to the control side not receiving aPTT. This result suggests that aPTT, as administered using ICG as the sensitizer and the small-scale LED-based light applicator surface, can help reduce gingivitis-associated local inflammation and provide additional therapeutic and putative preventive effects.

Greatest reduction of bacteria in dental plaque by aPTT was observed in streptococci indicating good potential in reduction of dental plaque cariogenicity. We found additional reduction in capnocytophaga, acinetobacteria, and rothia species. These low pathogenicity bacteria are generally associated with good health.^[27,28]^ We found the neisseria species to be relatively increasing in the plaque on the treated side. Neisseria have been associated with healthy oral flora, with a correlation to having a younger age, having a lower body mass index, having fewer caries, and being a non-smoker.^[29]^ Also, the haemophilus species were found to increase in the aPTT treated dental plaque. Haemophilus species have been shown in several studies to be endowed with healthy periodontal status,^[30]^ and larger amounts of haemophilus counts have been found in shallower periodontal pockets.^[31]^

In conclusion, our results demonstrate the feasibility of the self-care-compatible small-scale aPTT surface assembly and provide the first evidence of its clinical effectivity.

## Data Availability

The 16S rRNA sequences are deposited to the Sequence Read Archive (SRA) of the National Center for Biotechnology Information (NCBI) under BioProject ID PRJNA661546. Additional requests for data should be addressed to the corresponding author Dr. Tommi Patila

## ACKNOWLEDGEMENTS

We thank Nina Aspinen for help in dental photography, data collection, anonymization and analysis, and Saija Perovuo for technical assistance and support. We thank the personnel of DNA sequencing and genomics laboratory for making the 16S analysis work.

## AUTHOR CONTRIBUTIONS

T.P., S.N., J.R., A.K., N.M., J.M., E.K. and A.H. contributed to the design of the work. TS was responsible for the MMP-8 analysis, P.A. was responsible for the 16S bacterial analysis, SN was responsible for the plaque imaging, N.M. was responsible for the clinical study performance and sample collection under the guidance of A.K. All authors contributed to the drafting or revising the paper and contributed to the intellectual content of the work. All authors have approved the final version of the manuscript and ensure the integrity of the data collection and analysis for all aspects of the work.

